# Cardiological parameters predict mortality and cardiotoxicity in oncological patients

**DOI:** 10.1101/2023.05.05.23289440

**Authors:** Sebastian W. Romann, Daniel Finke, Markus B. Heckmann, Hauke Hund, Evangelos Giannitsis, Hugo A. Katus, Norbert Frey, Lorenz H. Lehmann

## Abstract

**Aims:** Oncological patients suspected to have the risk for cardiotoxicity, are recommended to be under intensified cardiological surveillance. We aimed to investigate the value of cardiac biomarker and patient-related risk factors (age, cardiovascular risk factors (CRF), cardiac function) for the prediction of all-cause mortality (ACM) and development of cardiotoxicity.

**Methods:** Between 01/2016 to 12/2020, patients with oncological diseases, admitted to the cardio-oncology unit at the University Hospital Heidelberg were included. They were examined by medical history, physical examination, 12-lead-ECG, 2D-echocardiography and cardiac biomarkers (high sensitive Troponin T (hs-cTnT); N-terminal brain natriuretic peptide (NT-proBNP)). Primary endpoint was defined as ACM, secondary endpoint was defined as cardiotoxicity as defined by the European Society of Cardiology.

**Results:** From 1971 included patients, primary endpoint was reached by 490 patients (25.7%) with a median of 363.5 [IQR 121.8, 522.5] days after presentation. Hs-cTnT of ≥7 ng/L (OR 1.82, p < 0.001) and NT-proBNP (OR 1.98, p < 0.001) were independent predictors of ACM, while reduced LVEF was not associated with increased ACM (p=0.85). Secondary endpoint was reached by 182 patients (9.2%) within a median of 793.5 days [IQR 411.2, 1165.0]. Patients with multiple CRF (defined as high-risk, n=886) had an increased risk for cardiotoxicity (n=100/886, 11.3%; HR 1.57, p=0.004). They showed increased baseline values of hs-cTnT (OR 1.60; p=0.006) and NT-proBNP (OR 4.00, p<0.001) and had an increased risk for ACM (OR 1.43; p=0.031).

**Conclusions:** In cancer patients, accumulation of CRF predetermines cardiotoxicity while increased hs-cTnT levels and NT-proBNP associate with ACM. Therefore, less intense surveillance protocols might be justified in patients with low values of cardiac biomarker and absence of CRF.

## Introduction

For most oncological diagnosis, the survival rates significantly improved over the last decade. This was achieved due to early detection and personalized treatment options with novel therapies (1, 2). Cancer treatment related cardiotoxicity as well as common risk factors for cancer and cardiovascular disease lead to a high cardiovascular mortality and morbidity in this patient cohort (3, 4). Moreover, there is growing evidence that cardiovascular diseases itself can promote the occurrence of cancer and vice versa (5, 6). In general, cardiovascular events such as thrombosis, pulmonary embolism, heart failure, arrhythmia or pericardial effusion have higher incidences in patients suffering from cancer, due to the malignant disease itself or in response to its therapy (7). While there is a general agreement that these patients need an intense cardiological care, only limited data are available predictive parameters that need to be integrated in standard surveillance strategies (8).

According to the current guideline of the European society of cardiology (ESC) a major task is to identify cancer patients with an increased baseline cardiovascular risk before the start of cancer treatment and minimizing their risk for cardiotoxicity before and during oncologic therapy with a specific surveillance program (8-11). While there are data available on the use of a standardized cardio-oncology workup in specific cancer patient cohorts, such as patients that are planned for an anthracycline therapy (12), the predictive value in an unselected cancer patient cohort is less clear. Moreover, it remains unclear how to effectively identify high-risk and low-risk patients for increased mortality and/or cardiotoxicity. We and others have previously reported the effectiveness of cardiac biomarkers regarding risk stratification of cancer patients (13, 14).

In the herewith reported large cohort of cardio-oncologic patients hs-cTnT effectively predicts ACM and identifies patients with risk for cardiotoxicity. NT-proBNP serves as an additional indicator for increased risk. Moreover, accumulation of cardiovascular risk factors, such as age, diabetes mellitus, family history of coronary artery disease, obesity, smoking and dyslipidemia are sufficient to identify a high-risk cohort for the development of cardiotoxicity.

## Methods

### Patients

Between 01/2016 to 12/2020, we collected data of 4315 patient’s visits from 1971 patients with oncological disease that were admitted to the Heidelberg Cardio-Oncology Registry (HEartCORE). The study protocol was approved by the Ethics Committee of the Medical Faculty, University Heidelberg (S-286/2017, 390/2011). The investigation conforms with the principles outlined in the *Declaration of Helsinki*.

The cardio-oncology outpatient clinic is part of the standard-of-care practice at the national center for tumor diseases (NCT) in Heidelberg and is offered as a standardized cardiological assessment for unselected oncological patients (8). Admission is based on the considerations of the treating oncologists. Every patient was examined by medical history, physical examination, 12-lead-ECG and 2D echocardiography including global longitudinal strain (GLS), if technical applicable. Additionally, cardiac biomarkers (high sensitive Troponin T (hs-cTnT) and N-terminal brain natriuretic peptide (NT-proBNP)) were assessed. Patients were administered usually before the begin of a new chemotherapy (1167, 59.2%).

In 1860 patients, hs-cTnT was measured. These patients were included in the following analysis regarding cardiac biomarkers. Primary Endpoint was defined as death of any cause, secondary endpoint was defined as impairment in the systolic left ventricular function as a drop in LVEF of more than 10% or below 50% or a relative reduction in GLS>15% from baseline, according to current guidelines (15, 16). Patients with initial LVEF, lower than 50% at first admission were not counted as an event. Only the further drop of LVEF or GLS abnormalities during the follow-up was then counted as a positive event.

If no cardiac alterations were found at the first presentation, we monitored the patients every 12 weeks during the oncological therapy. Patients with no further planned chemotherapy were only examined once (13.6%, n=269). In case of a reduced LVEF and/or increase in cardiac biomarker (hs-cTnT, NT-proBNP) and/or symptoms of heart failure we assessed the patients more frequently, in general every 4 weeks and eventually started or increased existing heart failure therapy. Further cardiac assessment via cardiac computer tomography (CT), cardiac magnet resonance tomography (MRI) or cardiac catheterization were based on clinical presentation, echocardiographic parameters and cardiovascular risk factors according to the current guidelines for acute and chronic coronary syndrome and heart failure (16-18).

### Data acquisition

Patient specific data was extracted from electronic medical records including ECG, laboratory results, echocardiographic measurements, cardiac MRI/CT results and angiographic results using the cardiac Research Data Warehouse (RWH).

Measurement of hs-cTnT in plasma samples was performed using the Elecsys® Troponin T high sensitive hs-cTnT assay (Roche Diagnostics) on a cobas® e411 immunoassay analyzer in the central laboratory at Heidelberg University Hospital. On Cobas e411, limit of blank (LoB), Limit of detection (LoD), 10% coefficient of variation (CV) and 99^th^ percentile cut-off values for the hs-cTnT assay were 3 ng/L, 5 ng/L, 13 ng/L and 14 ng/L (19, 20). N□terminal pro brain□type natriuretic peptide was measured by the Stratus® CS Acute Care™ NT□proBNP assay (Siemens AG, Berlin and Munich, Germany). Outcome data including ACM were acquired from the Clinical Cancer Registry of the National Centre for Tumor Diseases (NCT) Heidelberg. From the obtained data, it was not possible to retrace the number of deaths due to cardiovascular events.

Echocardiography was performed on a General Electrics (GE) Vivid E9 machine. Images were acquired ECG-triggered with at least three beats per image. LVEF was measured by a physician who is experienced in echocardiography using the biplanar calculation (2- and 4-chamber view). GLS was determined by use of a vendor-dependent analysis software (GE). The endocardial surface was detected automatically with manual adjustments, if necessary.

### Statistical analysis

All data was collected and processed in R, version 4.0.5 using in-house-scripting. The packages *ggplot2* (version: 3.3.2), *ggpubr* (version 0.4.0) and *survminer* (version 0.4.7) were used to illustrate plots. Kaplan-Meier curves were generated with the *survival* package (version: 3.2-3), we also used *relsurv* (version 2.2-3). The observational range was set to 3 years. Patients with shorter follow-ups were censored at the drop out time. The logrank test was used to determine differences in survival. A p-value < 0.05 was considered significant. Some graphs were illustrated in GraphPad Prism, version 7.4. In order to compare continuous variables, we used the Mann-Whitney Test. Dichotomic data was compared using the binomial distribution model. A confidence interval of 95% was considered significant. Univariate logistic regression (ulr) as well as multivariate logistic regression (mlr) analyses were performed in R.

Based on our previous data on potential cut-offs for hs-cTnT as a predictor of ACM in cancer patients (13), a hs-cTnT-threshold of 7ng/l was used for further analysis. Analyses regarding elevated hs-cTnT above the 99^th^ percentile (14 ng/l) (21) and above the cut-off regarding myocardial infarction above 52ng/l (18) can be found in **Suppl. Figure 2+3**. Elevated NT-proBNP were defined as levels above the age adjusted rule-out criteria of 450 pg/mL for <50 years, 900 pg/mL for 50-75 years and 1,800 pg/mL for >75 years (22). Median laboratory values in our cohort were used for further analysis to predict ACM. Patients were divided for risk assessment of cardiotoxicity into different risk groups according to age, cardiovascular risk factors and LVEF before therapy. Low risk of cardiotoxicity was defined for patients without cardiovascular risk factors and age between 18 and 50 years. Medium risk was defined for patients with one or two cardiovascular risk factors or age between 50 and 65 years. High risk for cardiotoxicity was defined for patients with more than two cardiovascular risk factors, age 65 years or older or LVEF<55% before therapy.

## Results

1971 patients with 4315 visits from before (n=1339), during (n=2126), after (n=705) or unspecified time point (n=145) regarding their systemic therapies between 01/2016 and 12/2020 were included into the analysis. In most cases (59.2%, n=1167) patients were examined before the beginning of a systemic therapy. 443 patients were initially presented in the cardio-oncology unit during chemotherapy (22.4%). Most of the patients were diagnosed with breast cancer (43.3%), upper gastrointestinal carcinoma (10.5%), multiple myeloma (6.0%) or melanoma (5.6%). The majority of patients was seen before or during an adjuvant or neoadjuvant setting (43.3%). 216 patients (11.3%) were admitted after terminated therapy or in palliative therapeutic regimes (448 patients, 22.7%). The first follow up was in median after 97 days (IQR 61, 147; n= 970) and the second follow up after another 90 days (IQR 57, 120; n=535). Follow-ups of the outcome were obtained for 543 days at median (IQR: 286.5, 928.5 days, n = 1903).

All patients received a cardiological assessment including medical history, physical examination, echocardiography including GLS and cardiac biomarkers (hs-cTnT, NT-proBNP), if applicable. Patients’ characteristics and demographics can be found in **Table 1**.

**Figure 5**
|  | Total | High-Risk | Medium-Risk | Low-Risk | p-value |
| --- | --- | --- | --- | --- | --- |
| **N** | 1971 | 886 | 840 | 245 |  |
| **Age [years]** | 60.5 [51.0, 69.2] | 69.7 [66.0, 75.5] | 55.9 [51.2, 60.5] | 39.9 [34.5, 46.4] | <0.001 |
| **Female gender** | 1277 (64.8%) | 503 (56.8%) | 572 (68.1%) | 202 (82.4%) | <0.001 |
| **Risk factors** |  |  |  |  |  |
| Smoking | 586 (31.1%) | 261 (30.7%) | 325 (40.3%) | 0 (0.0%) | <0.001 |
| Dyslipidaemia | 307 (16.3%) | 227 (26.7%) | 80 (9.9%) | 0 (0.0%) | <0.001 |
| Diabetes | 144 (7.4%) | 123 (14.2%) | 21 (2.5%) | 0 (0.0%) | <0.001 |
| Art. hypertension | 534 (27.6%) | 378 (43.5%) | 156 (18.9%) | 0 (0.0%) | <0.001 |
| Family history | 200 (10.3%) | 108 (12.4%) | 92 (11.1%) | 0 (0.0%) | <0.001 |
| Obesity | 149 (7.7%) | 103 (11.9%) | 46 (5.6%) | 0 (0.0%) | <0.001 |
| **Cancer diagnosis** |  |  |  |  |  |
| Breast cancer | 853 (43.3%) | 275 (31.0%) | 425 (50.6%) | 153 (62.4%) |  |
| Upper GI Tumor | 206 (10.5%) | 103 (11.6%) | 92 (11.0%) | 11 (4.5%) |  |
| Myeloma | 118 (6.0%) | 61 (6.9%) | 56 (6.7%) | 1 (0.4%) |  |
| Melanoma | 110 (5.6%) | 55 (6.2%) | 44 (5.2%) | 11 (4.5%) |  |
| Ovarial cancer | 54 (2.7%) | 27 (3.0%) | 20 (2.4%) | 7 (2.9%) |  |
| NET | 51 (2.6%) | 28 (3.2%) | 20 (2.4%) | 3 (1.2%) |  |
| Lymphoma | 101 (5.1%) | 57 (6.4%) | 33 (3.9%) | 11 (4.5%) |  |
| Leukaemia | 25 (1.3%) | 14 (1.6%) | 11 (1.3%) | 0 (0.0%) |  |
| Sarkoma | 80 (4.1%) | 31 (3.5%) | 34 (4.0%) | 15 (6.1%) |  |
| Lower GI | 43 (2.2%) | 28 (3.2%) | 12 (1.4%) | 3 (1.2%) |  |
| Lungen-CA | 44 (2.2%) | 38 (4.3%) | 5 (0.6%) | 1 (0.4%) |  |
| Other | 260 (13.2%) | 153 (17.3%) | 81 (9.6%) | 26 (10.6%) |  |
| **Oncologic therapy** |  |  |  |  |  |
| Curative | 223 (11.3%) | 119 (13.4%) | 87 (10.4%) | 17 (6.9%) |  |
| Palliative | 448 (22.7%) | 227 (25.6%) | 162 (19.3%) | 59 (24.1%) |  |
| Adjuvant | 324 (16.4%) | 131 (14.8%) | 147 (17.5%) | 46 (18.8%) |  |
| Neoadjuvant | 529 (26.8%) | 160 (18.1%) | 275 (32.7%) | 94 (38.4%) |  |
| Finished | 216 (11.0%) | 107 (12.1%) | 93 (11.1%) | 16 (6.5%) |  |
| **Number of visits** |  |  |  |  |  |
| 1 visit | 1971 (100%) | 886 (100%) | 840 (100%) | 245 (100%) |  |
| 2 visits | 970 (49.2%) | 394 (44.5%) | 439 (52.3%) | 137 (55.9%) |  |
| 3 visits | 535 (27.1%) | 210 (23.7%) | 257 (30.6%) | 68 (27.8%) |  |
| **Clinical chemistry** |  |  |  |  |  |
| Hs-TnT [ng/l] | 8 [5, 13] | 11 [7, 19] | 6 [4, 9] | 5 [3, 6] | <0.001 |
| NT-proBNP [ng/l] | 148 [76, 354] | 251 [129, 755] | 111 [61, 214] | 93 [54, 143] | <0.001 |
| Creatinine [mg/dl] | 0.74 [0.64, 0.88] | 0.79 [0.67, 1.02] | 0.72 [0.63, 0.84] | 0.66 [0.61, 0.74] | <0.001 |
| **Echocardiography** |  |  |  |  |  |
| LVEF | 59 [55, 60] | 56 [51, 60] | 60 [56, 60] | 60 [57, 60] | <0.001 |
| GLS [-%] | 18.3 [16.2, 20.4] | 17.5 [15.1, 19.9] | 18.9 [17.0, 20.6] | 19.1 [17.5, 21.2] | <0.001 |
| E/E' | 7.0 [5.0, 9.0] | 8.0 [6.0, 10.0] | 6.00 [5.0, 8.0] | 5.0 [4.0, 6.0] | <0.001 |
| **All-cause mortality** | 490 (25.7%) | 270 (31.7%) | 162 (20.0%) | 58 (24.3%) | <0.001 |
| **Cardiotoxicity** | 182 (9.2%) | 100 (11.3%) | 71 (8.5%) | 11 (4.5%) | 0.002 |

### Predictors for all-cause mortality in cancer patients

From all included patients, 490 patients died from any cause (25.7%) during the follow-up with a median survival of 363.5 [IQR 121.8, 522.5] days. Based on our previous data on potential cut-offs for hs-cTnT as a predictor of ACM in cancer patients (13), a hs-cTnT-threshold of 7ng/l was used for further analysis. A hs-cTnT of ≥7 ng/L (OR 1.82, p < 0.001) and NT-proBNP (OR 1.98, p < 0.001) were independent predictors of ACM among all included cardio-oncological patients (**Figure 1**) in multivariant analysis. Furthermore, hs-cTnT levels above the 99th percentile (14ng/l) showed a significantly higher ACM (OR: 2.07; p < 0.001) as well as they did for the cut-off regarding myocardial (18) infarction above 52ng/l (OR 1.94; p = 0.022, **Suppl. Figure 2+3**).

**Figure 1:**
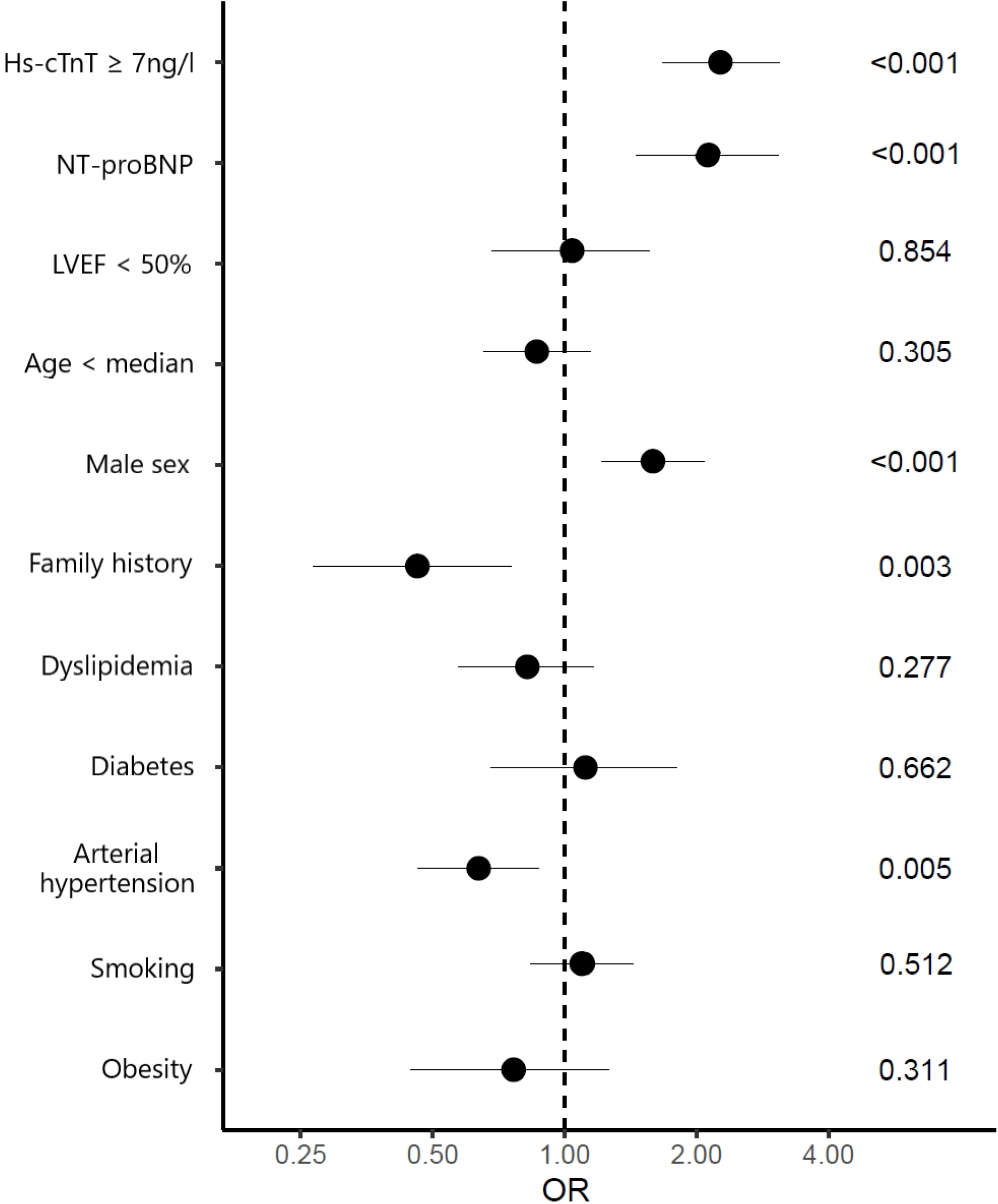
Multivariate logistic regression analysis on all-cause mortality (ACM). Odds rations (OR) and 95% confidence interval are shown as forrest blot. P-values as indicated. Hs-cTnT levels ≥ 7ng/l and age-adjusted NT-proBNP were associated with increased mortality. LVEF did show significant correlation only in univariate but not in multivariate analysis. Some tumor entities did significantly correlate with the outcome as well as arterial hypertension and a positive family history regarding cardio-vascular diseases. Hs-cTnT: high sensitivity cardiac troponin T; LVEF: left ventricular ejection fraction; NT-proBNP: N□terminal pro brain□type natriuretic peptide.

**Figure 2:**
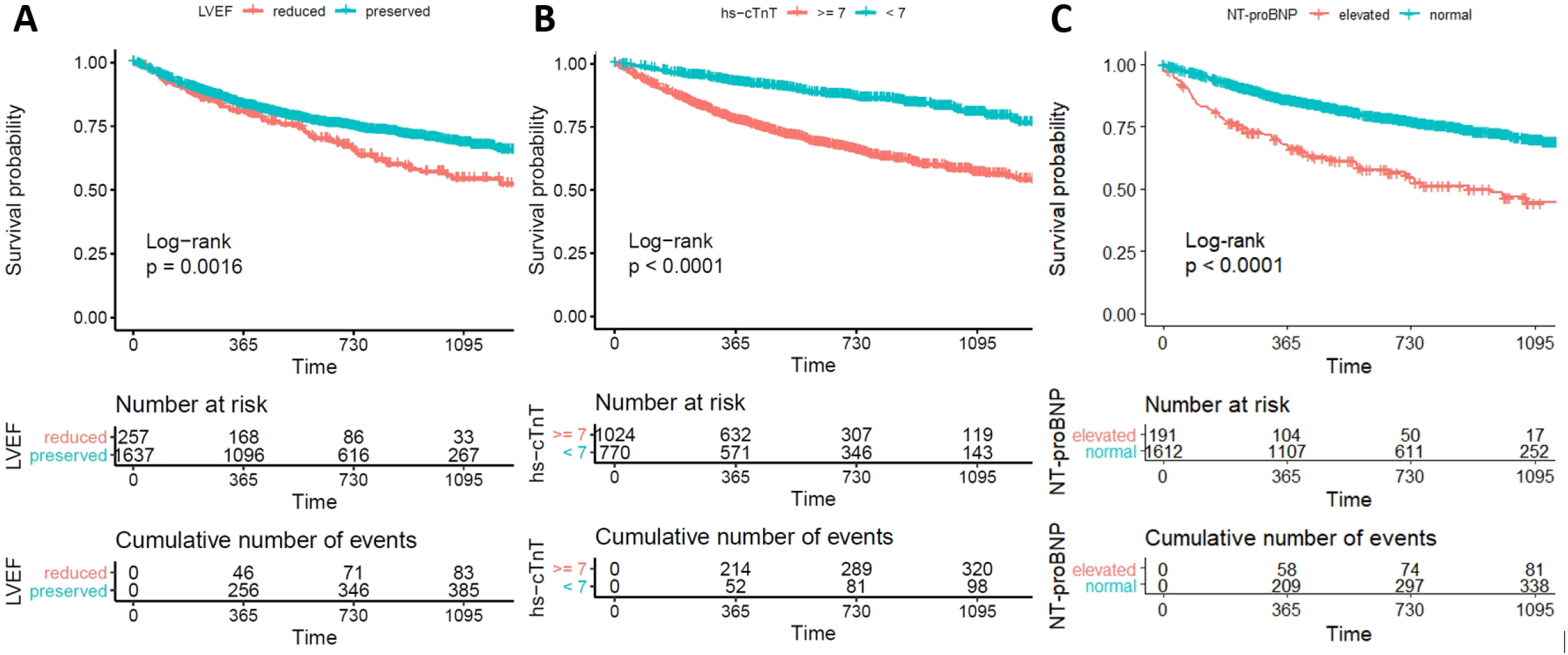
Kaplan Meier curves on all-cause mortality (ACM). Patients are divided into two groups according to their (A) left ventricular systolic function (LVEF ≥/< 50%), (B) NT-proBNP levels (age-adjusted clinical scale) and (C) hs-cTnT level (≥/< 7 ng/l). Logrank test p-value as indicated. Hs-cTnT: high sensitivity troponin T; LVEF: left ventricular ejection fraction; NT-proBNP: N-terminal brain natriuretic peptide.

Echocardiography showed in 90.6% of patients a preserved LVEF (>50%), with a slight decline at the first follow up after three months (87.3%, in median after 97 days [IQR 61, 147]) and the second follow up after six months (87.6%, in median after another 90 days [IQR 57, 120]) (**Suppl. Figure 1**). Univariate analysis of echocardiographic parameters reveals several statistical significant associations to ACM including LVEF reduction below 50% (OR 1.61; p=0.007), reduced Global longitudinal Strain (GLS>-18%; OR 1.89; p<0.001), increased posterior wall thickness above 10mm (OR 1.89; p<0.001) and septal wall thickness above 10mm (OR 1.36, p=0.006), increased left atrial size (OR 1.36; p=0.013), elevated vena cava inferior (OR 1.70; p=0.010) or pathological diastolic parameters E/A (OR 1.33; p=0.022) and E/e’ (OR 1.67; p=0.028) (**Figure 3**). However, LVEF <50% was not statistically significant associated with increased ACM in multivariate analysis (p=0.85) and failed as an independent predictor for ACM (**Figure 1**).

**Figure 3:**
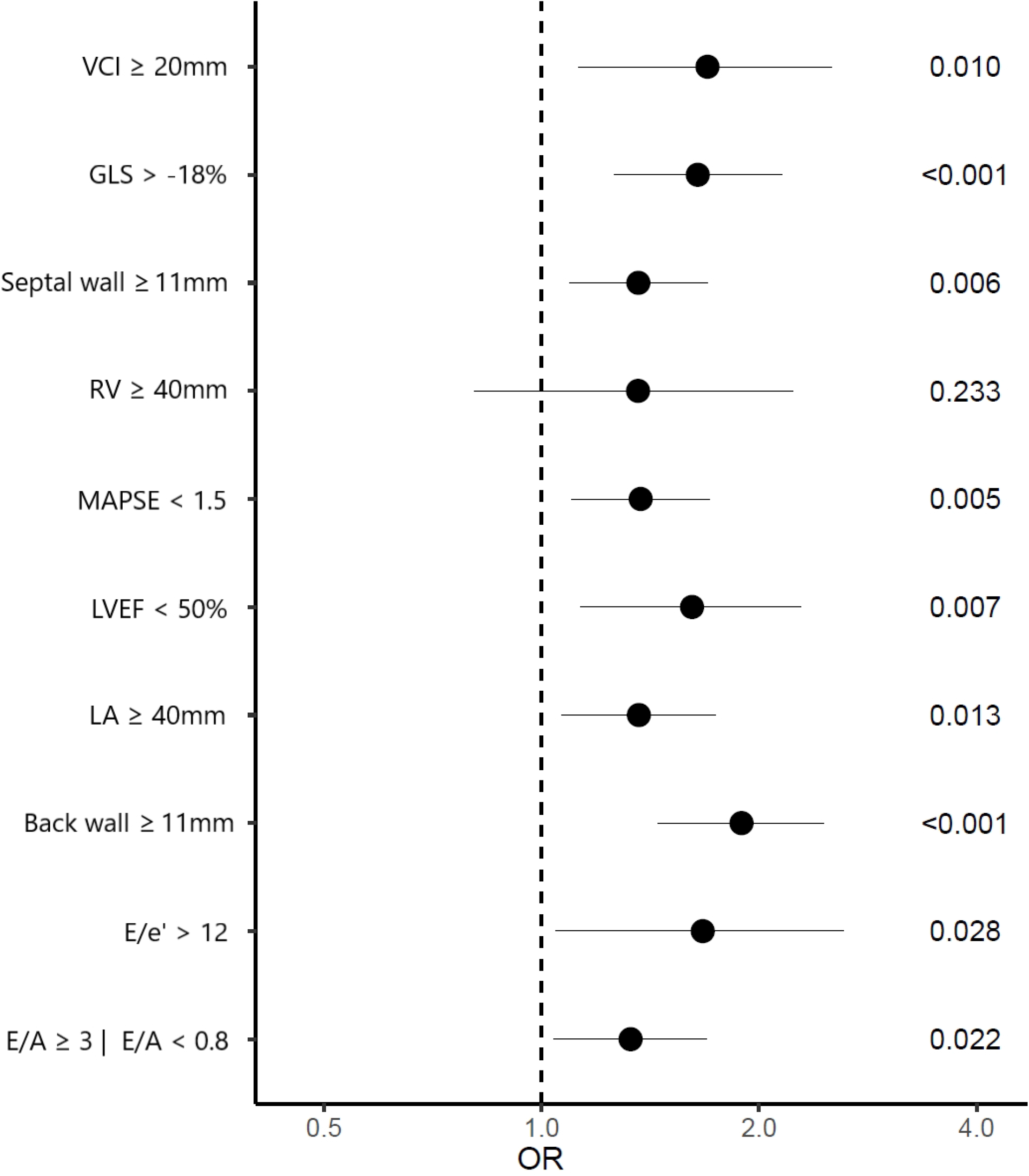
Univariate logistic regression analysis on all-cause mortality (ACM). Odds rations (OR) and 95% confidence interval are shown as forrest blot. Pathological LVEF, GLS, VCI, LA size, E/e’ and E/A scores as well as increased septal and posterior wall thickness were associated with increased mortality. P-values as indicated. VCI: Vena cava inferior size; GLS: global longitudinal strain; RV: right ventricular size; MAPSE: mitral annular plane systolic excursion; LVEF: left ventricular ejection fraction; LA: left atrial size; E/e’ and E/A: functional diastolic parameter.

### Predictors for cardiotoxicity

As a secondary endpoint, cardiotoxicity was defined according to the current guidelines of the ESC as a drop in LVEF of more than 10% and below 50% or a relative reduction in GLS>15% from baseline (15, 16). From all included patients, this endpoint was reached by 182 patients (9.2%) with a median follow-up time of 793.5 [IQR 411.2, 1165.0] days.

In univariate analysis, cardiotoxicity was significantly associated with increased baseline values of hs-cTnT (OR 1.60; p=0.006), age-adjusted NT-proBNP (OR 4.00, p<0.001) and presence of dyslipidemia (OR 1.52; p=0.035). Patients with cardiotoxicity had a significantly increased risk for ACM (OR 1.43; p=0.031) (**Figure 4**). Cardiotoxicity did not correlate significantly with the cardiovascular risk factors arterial hypertension (p=0.268), obesity (p=0.476), diabetes (p=0.846), positive family history (p=0.912) or smoking (p=0.422).

**Figure 4:**
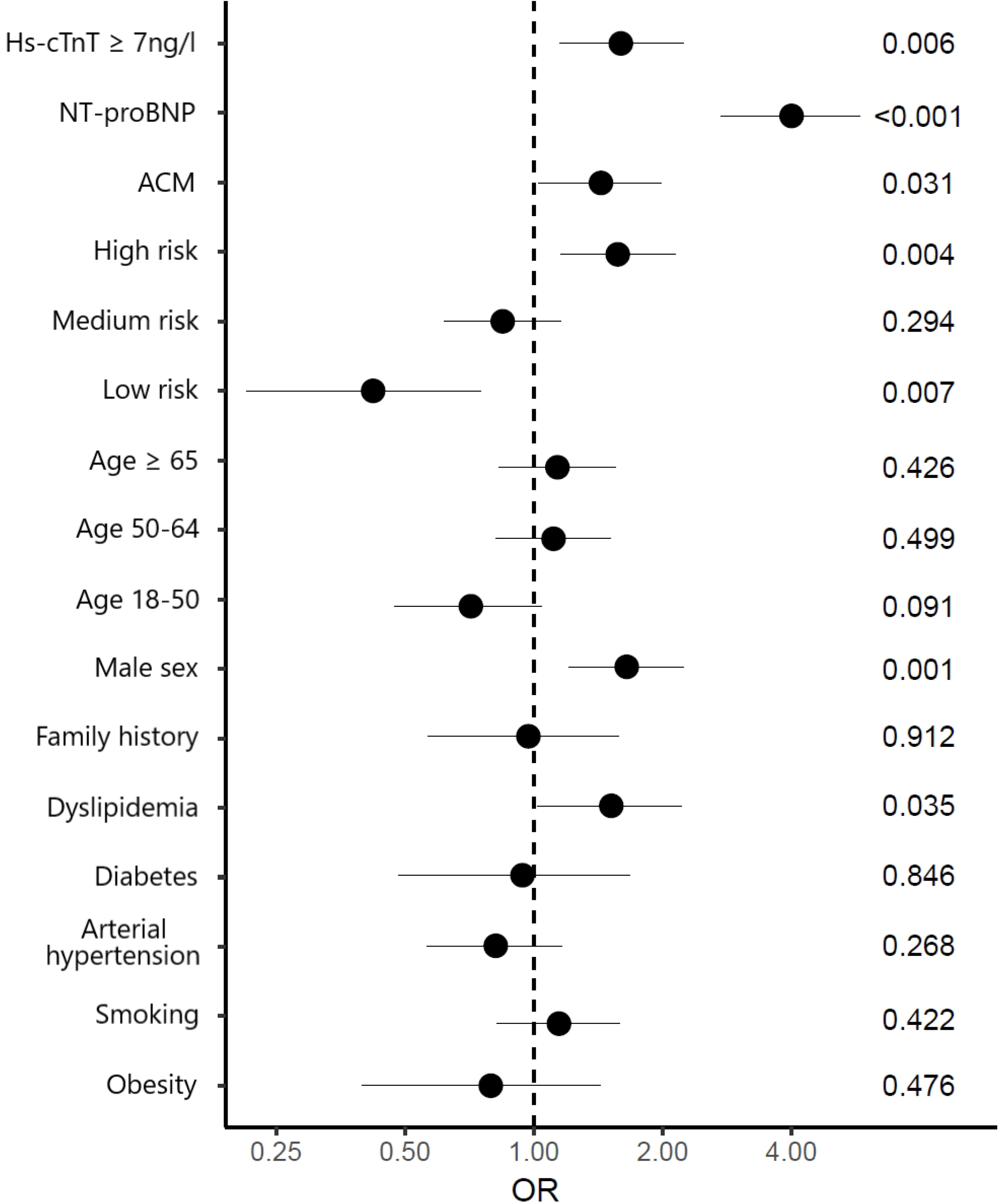
Univariate logistic regression analysis on Cardiotoxicity. Odds rations (OR) and 95% confidence interval are shown as forrest blot. ACM, elevated hs-cTnT levels ≥ 7ng/l and increased age-adjusted NT-proBNP values as well as high risk classification for cardiotoxicity, male gender and dyslipidemia were associated with increased Cardiotoxicity. Low risk for Cardiotoxicity (ESC) showed significant correlation with reduced risk for Cardiotoxicity. P-values as indicated. ACM: all-cause-mortality; Hs-cTnT: high sensitivity cardiac troponin T; NT-proBNP: N□terminal pro brain□type natriuretic peptide.

To identify a potential high-risk cohort for cardiotoxicity, patients were divided into different risk groups made up including the number of cardiovascular risk factors (**Figure 5A and B**). Accordingly, there were 12.4% patients in low risk group, 42.6% in medium and 45.0% in high risk group **(Figure 5C, Table 1**). Patients with high risk for cardiotoxicity (n=885) showed an increased number of cardiotoxic events (100 cases, 11.7%) after a median follow-up of 748 [IQR 408.2, 1161.0] days (HR 1.57, p=0.004). Low- and medium-risk cohort showed a younger and more healthy compilation of patients regarding cardiovascular risk factors. They were further characterized by more female patients as well as lower median levels of cardiac biomarkers. Details are summarized in **table 1**.

**Figure 5:**
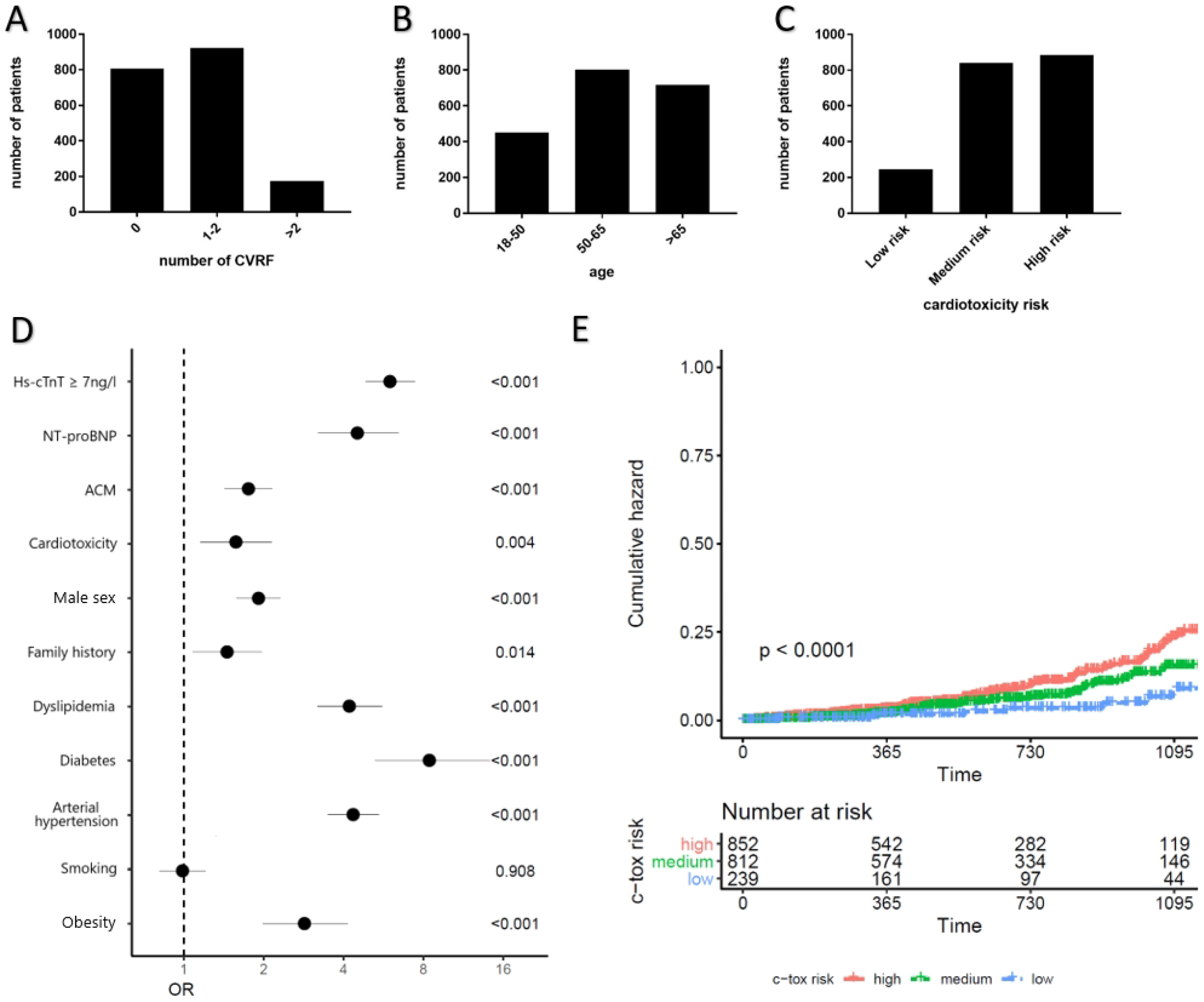
Number of patients regarding number of cardiovascular risk factors (CVRF; **A**), age (**B**) and cardiotoxicity risk groups regarding ESC (**C**). **D:** Univariate logistic regression analysis on high cardiotoxicity risk. Odds rations (OR) and 95% confidence interval are shown as forrest blot. ACM, male gender, reduced LVEF and elevated cardiac biomarkers as well as the CVRFs obesity, arterial hypertonia, diabetes, dyslipidemia and positive family history were associated with the high risk group. P-values as indicated. **E**: Cumulate hazard curves on cardiotoxicity. Patients are divided into groups according to their cardiotoxicity risk groups. Logrank test p-value as indicated. ACM: all-cause-mortality; Hs-cTnT: high sensitivity cardiac troponin T; LVEF: left ventricular ejection fraction; NT-proBNP: N□terminal pro brain□type natriuretic peptide.

### Cardiac risk factors identify a high-risk cohort per se

Univariate analysis (**Figure 5D**) showed significant correlations of the high risk group to ACM (OR 1.75; p<0.001), male gender (OR 1.91; p<0.001), reduced LVEF (OR 7.30; p<0.001) and elevated cardiac biomarkers (hs-cTnT: OR 5.99; p<0.001; NT-proBNP: OR 4.51; p<0.001), as well as the CVRFs obesity (OR 2.85; p<0.001), arterial hypertension (OR 4.35; p<0.001), diabetes (OR 8.43; p<0.001), dyslipidemia (OR 4.21; p<0.001) and positive family history (OR 1.46; p=0.014). Occurrence of cardiotoxicity was more frequent in patients with high-risk (**Figure 5E**). In line with an increased risk for cardiotoxicity, high-risk patients had a significantly higher mortality compared to low- and medium risk group (**suppl. Figure 4**).

## Discussion

Increasing survival rates of oncological patients have led to increased focus on the cardiovascular health of this patient cohort. In the five years’ follow-up from HEartCORE, a single-center registry for cardiac patients with underlying oncological therapy, our aim was to validate cardiac parameters stratifying these patients regarding their mortality and their cardiac health during oncological therapy. Most proposed factors for risk stratification hold true in this real-world patient cohort and support the notion of baseline risk assessment and diagnosis of cardiovascular disease as an important tool in cancer patients receiving cardiotoxic cancer treatments (11, 23). A meta-analysis of 61 trials showed a prediction of LV dysfunction in patients receiving cancer therapy by elevated troponin levels (24), and baseline troponin of cancer patients before chemotherapy is associated with ACM (13). This is particularly interesting, since patients with a low or very low hs-cTnT at baseline can be considered as low-risk oncological patients with regards to mortality. In case of therapies with low risk for cardiotoxicity, this group might be guided with less tight cardiological surveillance compared to patients with higher hs-cTnT levels. A comparison between the patients of our cutoff-defining study-cohort (13) which was analyzed taking the date of cancer diagnosis, and our cutoff-validation cohort can be found in the supplement **(Suppl. Figure 5**). In both cohorts, hs-cTnT separates low- and high-risk patients. The present study analyzed mortality from date of first admission in all patients. Notably, the predictive effect was not limited to ACM but also to the development of cardiotoxicity. In addition, age-adjusted increase in NT-proBNP-levels were significantly associated with ACM in cancer patients during cancer therapy and further underpin the impact of cardiac biomarkers during cardiological assessment and for further risk stratification.

Following the current guidelines of the European Society of Cardiology (ESC) the German Society of Cardiology (DGK) and practical guidelines of the American Society of Clinical Oncology (ASCO), echocardiographic parameters are one pillar of cardio-oncological risk stratification, including systolic and diastolic left ventricular function, as well as GLS analysis (1, 25, 26). Based on the recommendations, the general number of patients, theoretically eligible for intensive cardio-oncological assessment are high (e.g., all patients with mamma carcinoma at age >65 years). Therefore, sequential measuring of cardiac biomarker might be implemented as a gating tool for intensified cardio-oncological assessment. This would allow to reduce the total number of patients, where echocardiography is currently indicated.

### Patient-related risk factors for the identification of a high-risk cohort

As known from other cardiovascular diseases (17), pretest probability seems to allow a further risk-stratification beside measuring cardiac biomarker. Two classical risk factors for the development of atherosclerosis (dyslipidemia and male gender) were associated with an increased risk for cardiotoxicity. However, age, diabetes, family history of cardiovascular events, arterial hypertension and obesity were not independently associated with an increased risk for cardiotoxicity. To our knowledge, association between cardiac risk factors (presence of CAD, diabetes, age, hypertension and renal dysfunction) and increased risk for cardiotoxicity was only shown in older women after adjuvant trastuzumab therapy (27). Our patient cohort clearly differ from that, which might explain the less predictive value of patient-related risk factors.

While single risk factors were not independently associated with higher risk for cardiotoxicity, patients accumulating multiple risk factors showed and increased risk. Of note, this increased risk for cardiotoxicity was occurring after three years of follow-up which is in line with previous observations from trastuzumab-related cardiotoxicity (27). Therefore, our data support the notion that patients with accumulation of multiple cardiac risk factors have a high risk for the development of cardiotoxicity in the long-term follow-up and might justify an intensified cardio-oncological surveillance protocol, as it is suggested by the current ESC position paper (11, 28). This patient cohort was additionally characterized by a slightly increased hs-cTnT level at baseline, supporting the predictive role of hs-cTnT in the identification of high-risk patients.

## Conclusions

Measurement of cardiac hs-cTnT can possibly be used for the risk stratification (for ACM and development of cardiotoxicity) of cancer patients. Especially hs-cTnT values which were measured before initializing a chemotherapy were able to identify patients at high-risk. Patients with higher baseline hs-cTnT plasma concentration (≥ 7ng/l) should be considered for a tighter cardiac surveillance strategy. Hs-cTnT might additionally be implemented in cancer studies to early identify high-risk patients. Beside cardiac biomarker, accumulation of multiple cardiac risk factors (>2 and/or age >65) identify a cohort with increased ACM and higher risk for cardiotoxicity.

Further prospective, randomized, multi-center trials need to confirm these findings in other clinical cohorts. It further needs to be determined, whether functional measurements including echocardiography might be dispensable in low-risk patients characterized by hs-cTnT levels below 7ng/l.

### Competencies in Medical Knowledge

In oncological patients, cardiological assessment is frequently requested if risk for cardiotoxicity is considered. Patients with low values of cardiac biomarker and limited number of classical cardiovascular risk factors may be considered as a low-risk cohort. In these patients a less frequent surveillance might be justified.

### Study limitations

Patients included into the study are heterogeneous based on different cancer entities and different anti-cancer therapies. This is a real-world collective without patient pre-selection according to specific oncological or cardiological parameters. Patients were included before, during and after their oncologic therapy. During follow-up consultations, there is a possible selection bias towards patients with pathological findings (e.g. elevated cardiac biomarker or reduction of LVEF) that were admitted to a more stringent follow-up regime. However, we have largely standardized a follow-up of three month in all cancer patients during therapy. Patients who presented after a terminated therapy can be considered as cancer survivors and showed a considerably lower overall mortality rate. Data regarding ACM were retrieved from the Clinical Cancer Registry of the National Centre for Tumor Diseases (NCT) Heidelberg. The causes of death could not be linked to specific pathologies. Secondary endpoint was defined as decline in LVEF during treatment. Certain forms of CV toxicity may be present, including vascular/ischemic-, thrombotic-, arrhythmic-side effect or HFpEF, which might not be captured by the decline in LVEF (29).

## Data Availability

All data produced in the present study are available upon reasonable request to the authors

## Acknowledgements

We thank Monika Arnold and Ines Ludwig for support with clinical data collection and patient care.

## Data Availability Statement

Data available on request.

