## Supplementary figures and images for "Cardiological parameters predict mortality and cardiotoxicity in oncological patients"

### Supplement

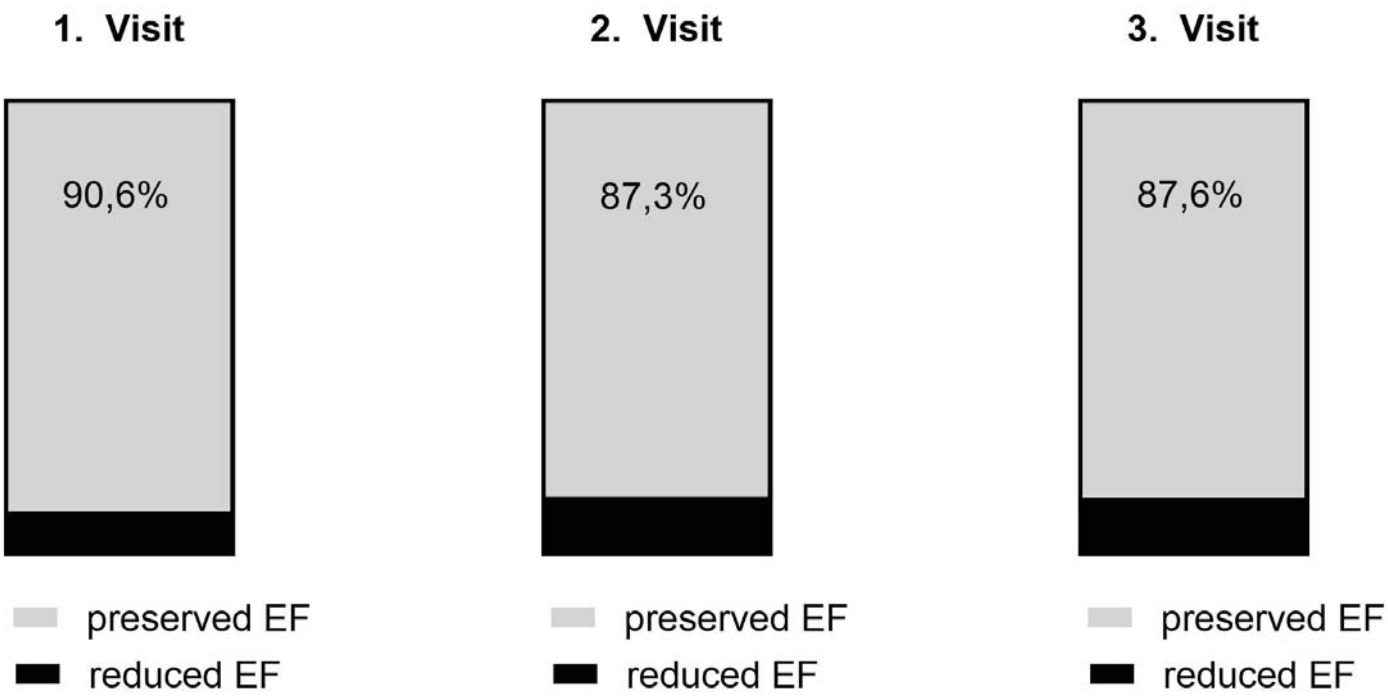


**Suppl. Figure 1**

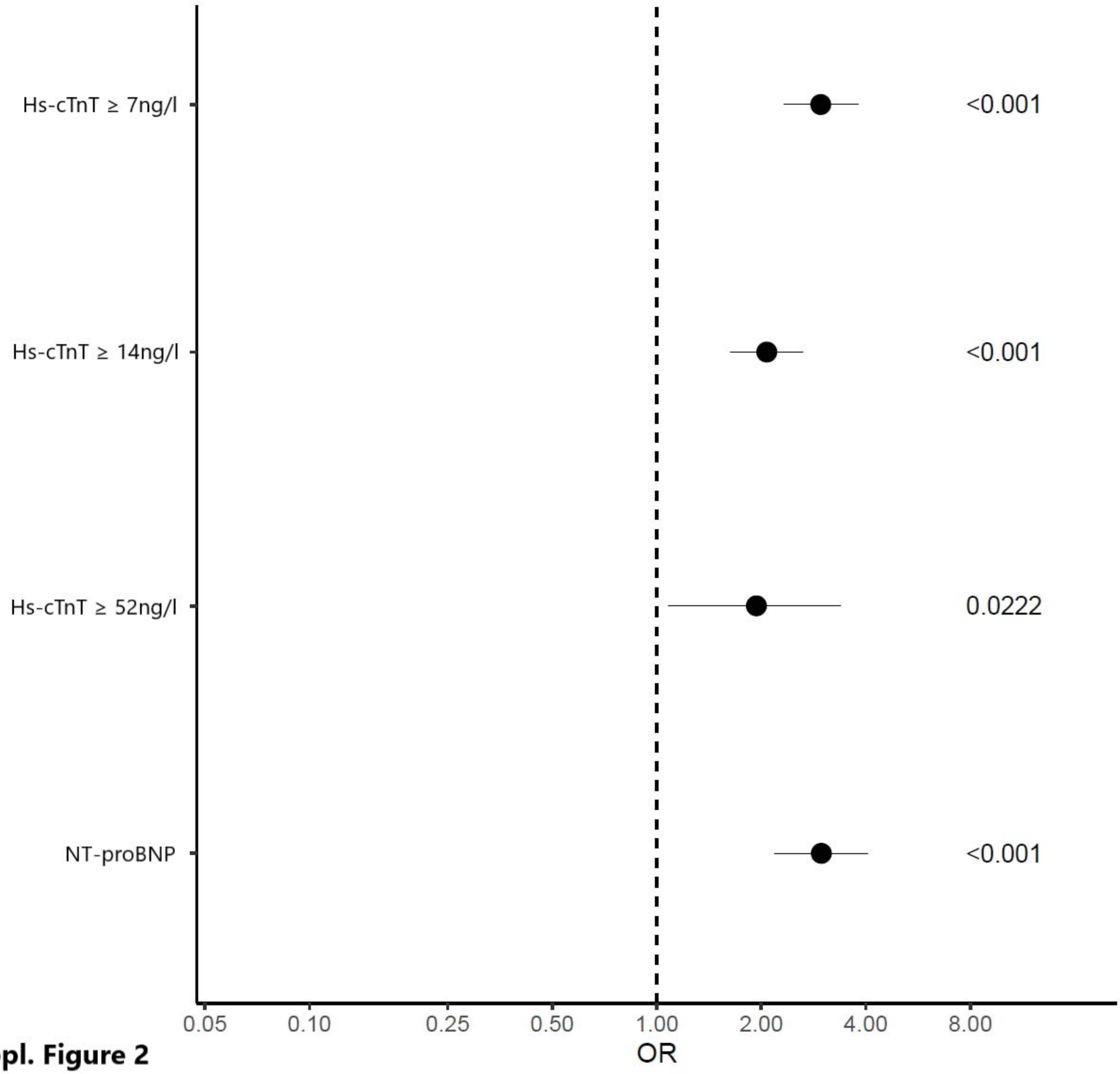


**Suppl. Figure 2**

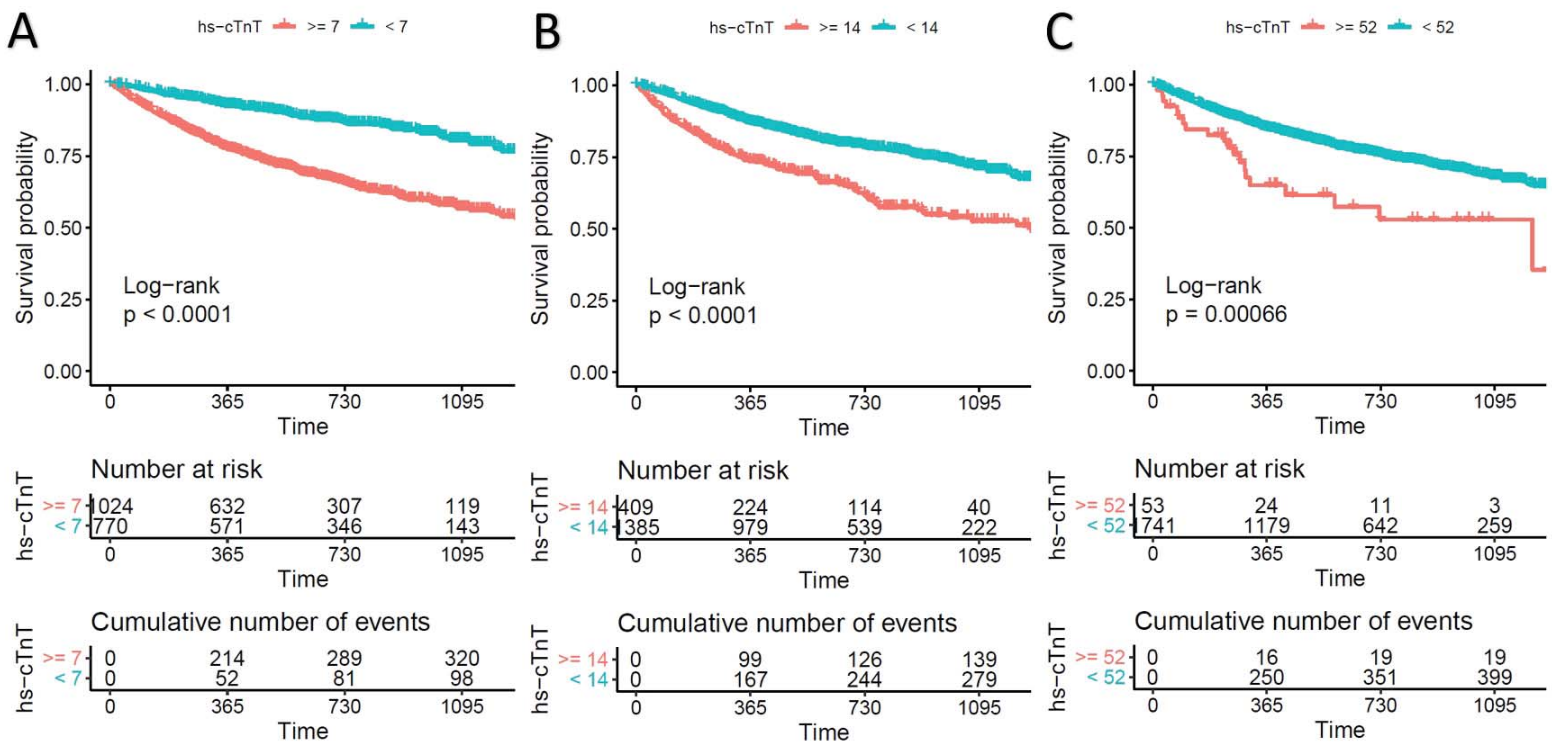


**Suppl. Figure 3**

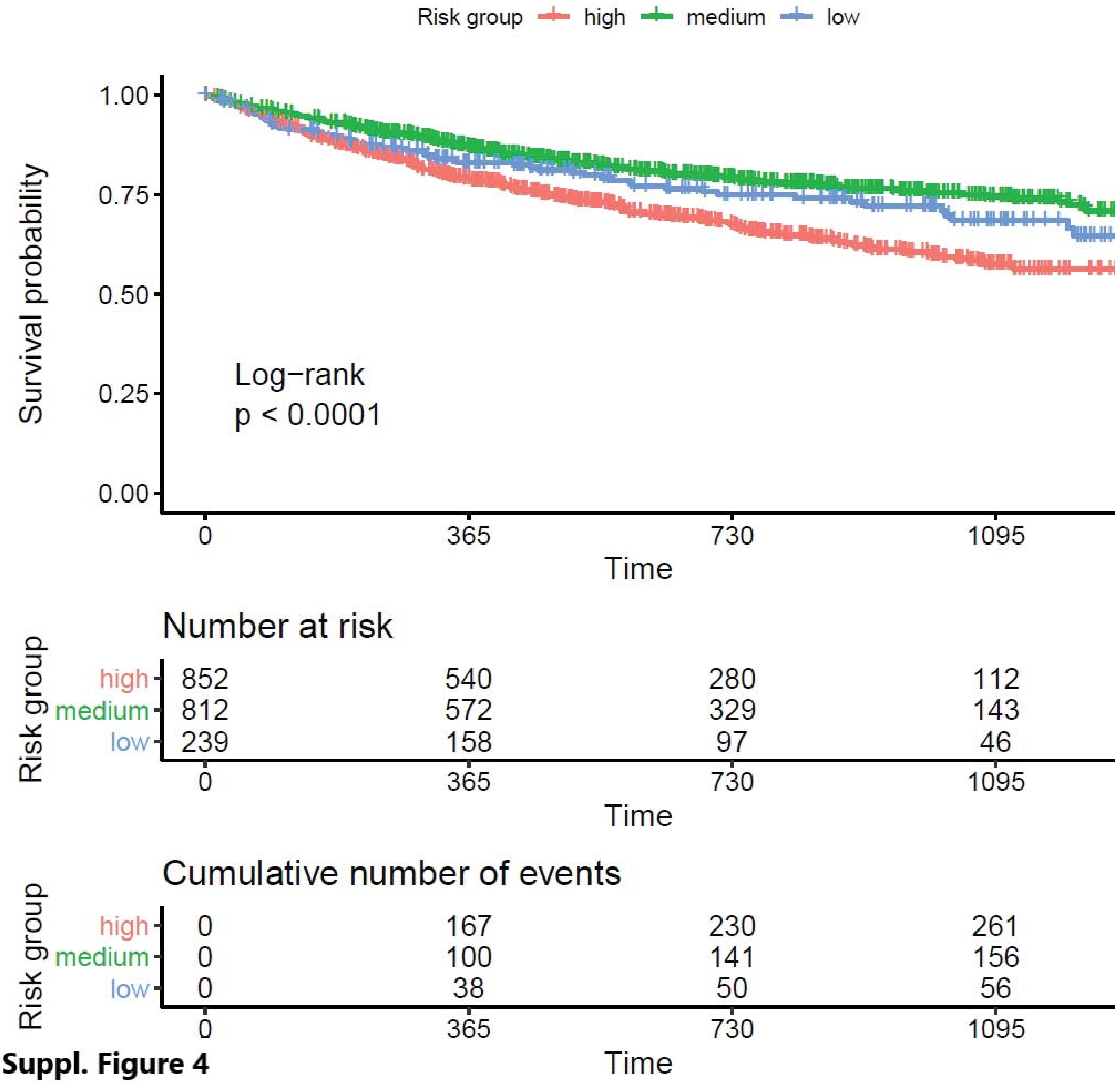


**Suppl. Figure 4**

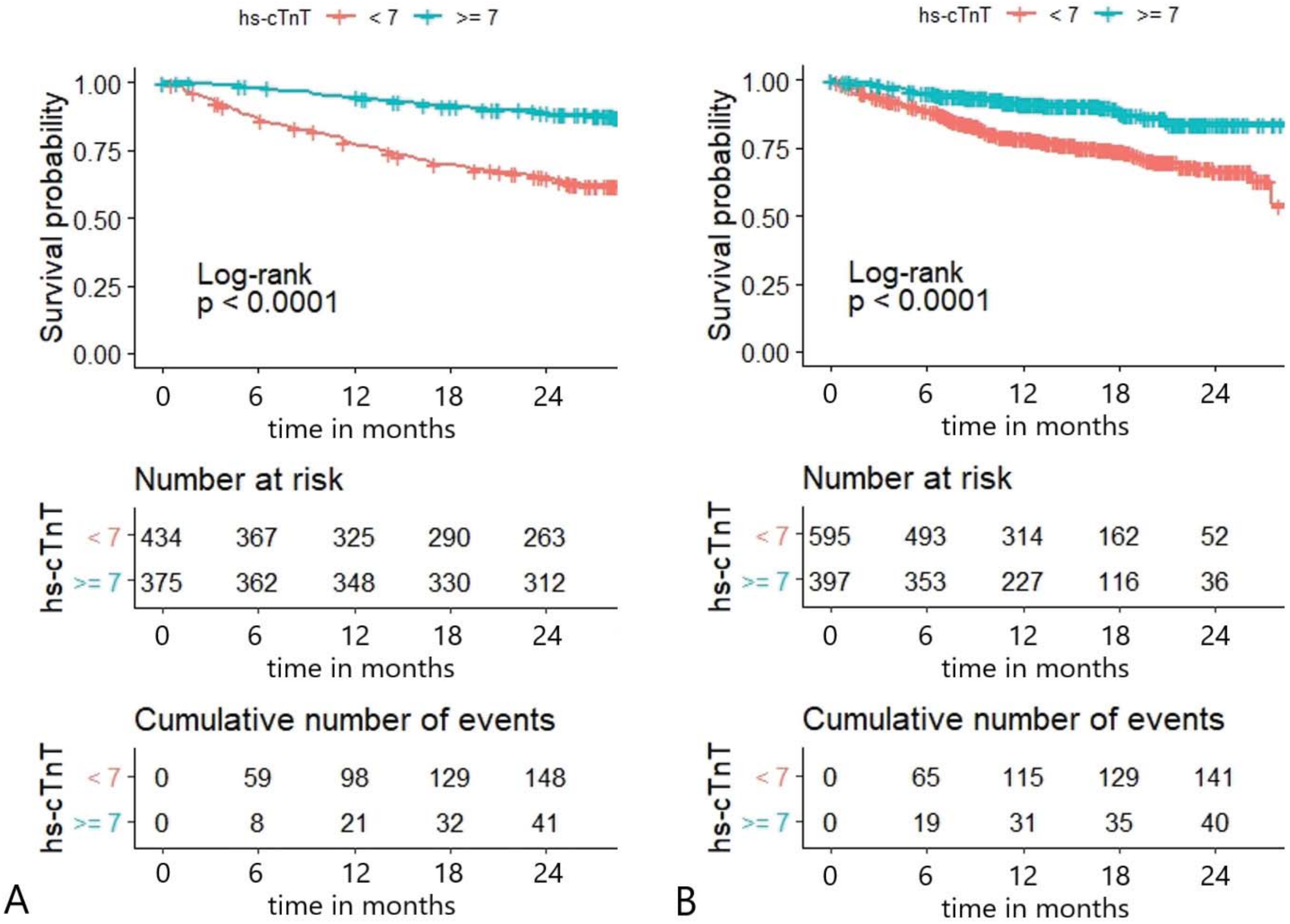


**Suppl. Figure 5**
